# Estimation of positron-emission-tomography amyloid load and related biomarkers in Alzheimer’s disease using evoked potential tomography electroencephalography

**DOI:** 10.1101/2025.07.08.25331158

**Authors:** Boris-Stephan Rauchmann, James Hamet, Jesyin Lai, Homeira Kafi, Joe Rexwinkle, Matthias Brendel, Nicolai Franzmeier, Carolin Kurz, Oliver Pogarell, Johannes Levin, Günter Höglinger, Robert Perneczky

## Abstract

**INTRODUCTION:** Dementia affects over 50 million individuals globally, predominantly due to Alzheimer’s disease (AD). Effective early detection, monitoring, and intervention remain clinical challenges, and existing diagnostic frameworks lack unified portable solutions for assessing multiple biomarkers.

**METHODS:** This study evaluates Evoked Potential Tomography (EPT), an EEG-based technique using a novel visual evoked potential protocol. We developed an automated pipeline including EEG preprocessing, event-related potential feature extraction, feature selection, optimization, and regression modeling to estimate key AD biomarkers: PET-amyloid standardized uptake value ratio (SUVR), CSF phosphorylated tau 181 (*p*-tau181), Free and Cued Selective Reminding Test (FCSRT) scores, and Mini-Mental State Examination (MMSE) scores.

**RESULTS:** Regression modeling of ERP features from early-AD participants yielded strong Spearman’s correlations between predicted and true values for PET-amyloid SUVR, *p*-tau181, FCSRT, and MMSE scores, with r = 0.8-0.94.

**DISCUSSION:** Despite limitations, findings support EPT’s potential for sensitive, accurate, and non-invasive early AD detection and progression monitoring. Further validation is ongoing.

**Highlights:**

- ERP amplitudes differed by dementia diagnosis during visual stimulation.
- ERP regression outperformed CSF amyloid ratio in PET amyloid estimation.
- ERP features predicted *p*-tau and cognition (r = 0.8–0.94).
- Evoked Potential Tomography may track Alzheimer’s progression effectively.

**Research in Context:**

1. **Systematic Review:** The authors conducted a systematic literature review using traditional databases (e.g., PubMed). Although Alzheimer’s disease (AD) biomarkers are extensively studied, no single portable technology currently assesses all essential biomarkers simultaneously. Relevant references are appropriately cited.
2. **Interpretation:** Our results demonstrate that integrating the Evoked Potential Tomography (EPT) protocol with an automated EEG processing and biomarker estimation pipeline effectively estimates multiple critical AD biomarkers that are useful in tracking disease progression using a single 30-minute EEG session.
3. **Future Directions:** The study highlights the promise of EPT as a non-invasive tool for AD screening and diagnostics. Further validation of this method could significantly advance clinical management and research, facilitating earlier AD detection, precise cohort selection, and targeted therapeutic interventions.

## Introduction

Analyses of brain tissue from human and animal models suggest that fibrillar amyloid-β (Aβ) negatively impacts both excitatory neurons and inhibitory terminals, disrupting neuronal function and causing network-level damage [1,2]. Research involving transgenic mice with Alzheimer’s Disease (AD) overexpressing Aβ demonstrates that neuronal hyperactivity in cortical and hippocampal regions, disruption of slow-wave oscillations, and network hypersynchrony precede plaque formation, indicating that hyperactivity may be one of the earliest physiological disturbances in AD pathogenesis [3,4]. Additionally, phosphorylated tau (*p*-tau) impairs microtubule structure in axons both *in vitro* and in mice [5], resulting in a progressive reduction in synaptic quantity and efficiency, and disrupting intra- and inter-regional brain communication [6]. Moreover, recent findings suggest that cerebrospinal fluid (CSF) *p*-tau significantly modulates neuronal excitability and network activity in AD and related tauopathies [7].

Alterations in neural activity observed in animal models can now be examined in humans through electroencephalography (EEG), particularly through resting-state functional brain network analysis and event-related potential (ERP) analyses. These methods reveal changes in functional connectivity even in healthy aging individuals [8] and prodromal AD cases [9]. Despite advancements in connectivity analysis, EEG remains underutilized in standard AD diagnostics. Given the cost-effectiveness, non-invasiveness, and widespread availability of EEG equipment in clinical settings, EEG analysis represents a promising tool for population-wide AD screening and early risk detection. Previous studies have shown correlations between CSF Aβ status and EEG connectivity measures in cognitively normal older adults [10]. Additionally, patients with mild cognitive impairment (MCI) who are CSF Aβ-positive exhibit increased global slowing in brain oscillatory activity compared to Aβ-negative controls [11]. Evidence also links tau pathology to network disruptions detected by magnetic resonance imaging (MRI) in fully symptomatic AD [12] and other neurodegenerative diseases like progressive supranuclear palsy [13]. Recent developments in machine learning and artificial intelligence have facilitated automated assessments of EEG data, enabling methods that simultaneously learn meaningful features from the data and classify disease states [10].

Furthermore, recent research has demonstrated that EEG analysis can achieve high accuracy in distinguishing individuals at risk for AD from healthy controls [14]. Novel approaches include connectivity measures such as coherence or phase lag and graph analysis characteristics combined with data-driven machine learning and deep learning models, which enhance the detection and classification of neurological and mental health disorders [10,15,16]. Independent studies report that individuals with subjective cognitive decline, the earliest clinical stage of AD, who are positive for Aβ, exhibit enhanced functional resting-state connectivity in the alpha frequency band correlated with higher global Aβ load and reduced connectivity in the beta frequency band [17,18]. However, the direct association between early electrophysiological changes and pathological hallmarks of AD remains to be firmly established through rigorous statistical and machine learning methodologies.

In addition to pathological biomarkers such as amyloid and *p*-tau, clinical cognitive assessments are essential tools in evaluating memory and overall cognitive functioning in AD research. Among these assessments, the Free and Cued Selective Reminding Test (FCSRT) and the Mini-Mental State Examination (MMSE) are widely utilized due to their sensitivity and practicality. The FCSRT specifically assesses verbal episodic memory and has proven particularly useful in characterizing memory impairments within the AD spectrum [19]. For instance, patients exhibiting lower FCSRT free recall scores demonstrate higher prevalence rates of dementia and an increased risk of progressing to dementia in the future [20] Complementing the FCSRT, the MMSE is frequently employed as a screening tool for dementia, predicting cognitive decline [21], and monitoring cognitive changes over time [22].

Thus, the primary aim of this study was to investigate the associations between visually evoked responses captured by EEG and key pathological changes and clinical cognitive scores in individuals with AD, MCI, and age-matched controls within a prospective clinical study. We utilized a novel Evoked Potential Tomography (EPT) protocol comprising optimized visual stimuli that systematically varied in contrast, spatial frequency, and relative movement to selectively activate distinct neuronal populations involved in early visual processing. A similar protocol has previously been shown to effectively differentiate between patients with MCI and AD, revealing distinct spectral features correlated with continuous PET-amyloid SUVR [23]. Moreover, this protocol has demonstrated sensitivity to cognitive variations in schizophrenia and autism spectrum disorders [24,25], highlighting its potential as a broadly applicable tool for assessing neural processing integrity. Consequently, we hypothesized that EEG features extracted during EPT could accurately estimate PET-amyloid SUVR, *CSF p*-tau181 values, and other cognitive scores, such as FCSRT free recall and MMSE, across individuals at varying stages of dementia.

## METHODS

### Study design and participants

The data used in this study originate from the baseline dataset of the ActiGliA study, a prospective, longitudinal observational study within the Munich Cluster for Systems Neurology (SyNergy) at Ludwig-Maximilians-University (LMU) Munich, initiated in 2017 [26]. Participants analyzed in this study were recruited through a specialized outpatient clinic at the LMU hospital Department of Psychiatry and Psychotherapy. Participants were included after providing written informed consent according to the Declaration of Helsinki. The study was approved by the ethics committee of LMU Munich (project numbers 17-755 and 17-569). ActiGliA includes extensive assessments that cover neurocognitive evaluations, clinical examinations, MRI and PET imaging, as well as bio-banking of fluids, including CSF testing. Further details on the study design, participant characteristics, and methods are described elsewhere [26]. Out of 124 ActiGliA participants, 73 met the inclusion criteria, with EEG recordings available for 48 of these individuals. This resulted in a cohort consisting of 16 participants with AD, 8 participants with MCI (mild dementia), 22 age-matched control participants with close-to-normal neurodegeneration CSF biomarkers (amyloid and *p*-tau), and 2 participants with non-Alzheimer’s dementia (NAD). Demographic information for these participants is summarized in Table 1.

**Table 1:**
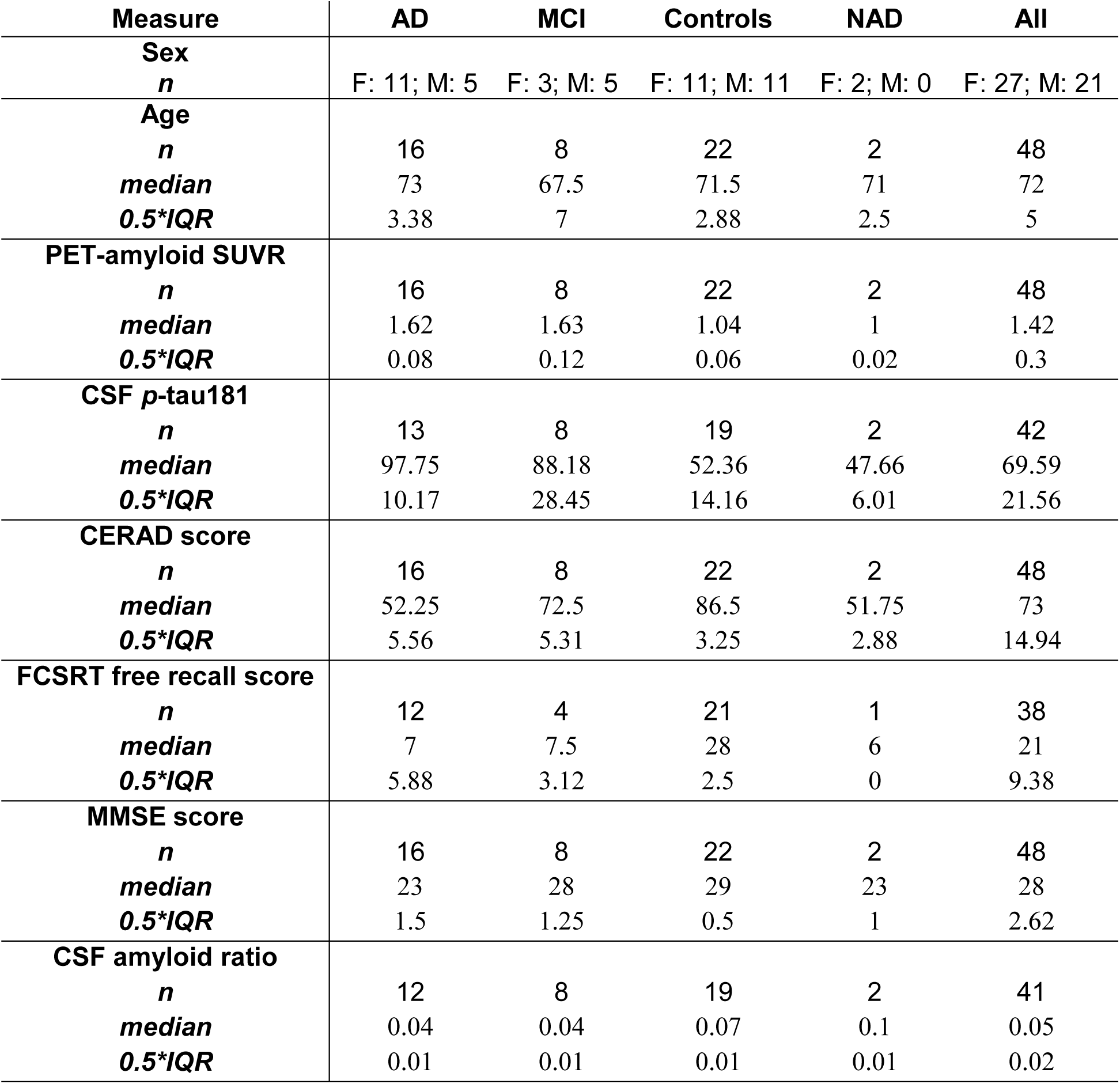
Demographic info of participants with EEG recordings. AD = Alzheimer’s disease; NAD = non-Alzheimer’s dementia; PET = positron-emission tomography; SUVR = standardized uptake value ratio; *p*-tau = Phosphorylated tau; CERAD = Consortium to Establish a Registry for Alzheimer’s Disease Neuropsychological Battery; FCSRT = Free and Cued Selective Reminding Test; MMSE = Mini-Mental State Examination; IQR = inter-quartile range.

| Measure | AD | MCI | Controls | NAD | All |
| --- | --- | --- | --- | --- | --- |
| **Sex** |  |  |  |  |  |
| *n* | F: 11; M: 5 | F: 3; M: 5 | F: 11; M: 11 | F: 2; M: 0 | F: 27; M: 21 |
| **Age** |  |  |  |  |  |
| *n* | 16 | 8 | 22 | 2 | 48 |
| *median* | 73 | 67.5 | 71.5 | 71 | 72 |
| *0.5\*IQR* | 3.38 | 7 | 2.88 | 2.5 | 5 |
| **PET-amyloid SUVR** |  |  |  |  |  |
| *n* | 16 | 8 | 22 | 2 | 48 |
| *median* | 1.62 | 1.63 | 1.04 | 1 | 1.42 |
| *0.5\*IQR* | 0.08 | 0.12 | 0.06 | 0.02 | 0.3 |
| **CSF *p*-tau181** |  |  |  |  |  |
| *n* | 13 | 8 | 19 | 2 | 42 |
| *median* | 97.75 | 88.18 | 52.36 | 47.66 | 69.59 |
| *0.5\*IQR* | 10.17 | 28.45 | 14.16 | 6.01 | 21.56 |
| **CERAD score** |  |  |  |  |  |
| *n* | 16 | 8 | 22 | 2 | 48 |
| *median* | 52.25 | 72.5 | 86.5 | 51.75 | 73 |
| *0.5\*IQR* | 5.56 | 5.31 | 3.25 | 2.88 | 14.94 |
| **FCSRT free recall score** |  |  |  |  |  |
| *n* | 12 | 4 | 21 | 1 | 38 |
| *median* | 7 | 7.5 | 28 | 6 | 21 |
| *0.5\*IQR* | 5.88 | 3.12 | 2.5 | 0 | 9.38 |
| **MMSE score** |  |  |  |  |  |
| *n* | 16 | 8 | 22 | 2 | 48 |
| *median* | 23 | 28 | 29 | 23 | 28 |
| *0.5\*IQR* | 1.5 | 1.25 | 0.5 | 1 | 2.62 |
| **CSF amyloid ratio** |  |  |  |  |  |
| *n* | 12 | 8 | 19 | 2 | 41 |
| *median* | 0.04 | 0.04 | 0.07 | 0.1 | 0.05 |
| *0.5\*IQR* | 0.01 | 0.01 | 0.01 | 0.01 | 0.02 |

### Clinical assessments

The Clinical Dementia Rating and the Consortium to Establish a Registry for Alzheimer’s Disease Neuropsychological Battery (CERAD) [27] were administered by trained psychologists at the LMU hospital memory clinic. A CERAD total score was computed following methods previously described by Chandler et al. (2005) [28], integrating results from six cognitive subtests: semantic fluency (number of animals named in 60 seconds), modified Boston Naming Test, Word List Learning, Constructional Praxis, Word List Recall, and Word List Recognition Discriminability. Additionally, participants completed the FCSRT, a memory test specifically designed to assess verbal episodic memory [29], as well as the MMSE, which evaluates orientation, memory, attention, and language [30,31]. The free recall component of the FCSRT is especially sensitive in detecting memory impairments associated with MCI and AD [20]. Together, higher scores on the CERAD battery, FCSRT free recall, and MMSE collectively indicate better cognitive functioning, providing a comprehensive picture of cognitive status alongside neuropathological assessments.

### Cerebrospinal fluid analyses

CSF peptide measures were obtained from aliquoted samples using commercially available enzyme-linked immunosorbent assays (ELISAs; Fujirebio, Malvern, PA). Aβ positivity was defined as a CSF Aβ42/40 ratio of less than 5.5%, following previously established criteria [32]. The concentrations of total tau and *p*-tau181 were measured using Innotest htau-Ag and Innotest *p*-tau ELISA assays (Fujirebio, Europe).

### EEG recordings and stimulus paradigm

Neural signals were recorded using a 64-channel Waveguard EEG headset (Ag/AgCl electrodes) arranged equidistantly and connected to an ANT recording system (ANT Neuro, Enschede), sampled at 2048 Hz. Participants completed a visual stimulation paradigm [23,25] comprising a series of distinct visual stimulus types designed to concurrently evaluate various aspects of early visual processing (Fig. 1). Visual stimuli were displayed against an isochromatic grey background (RGB: 127, 127, 127). Participants were instructed to maintain fixation on a central cross and to ignore the visual stimulus as it was presented. To maintain participant attention, participants were instructed to press the space bar when the fixation cross dimmed slightly (every 3–12 seconds).

**Figure 1.**
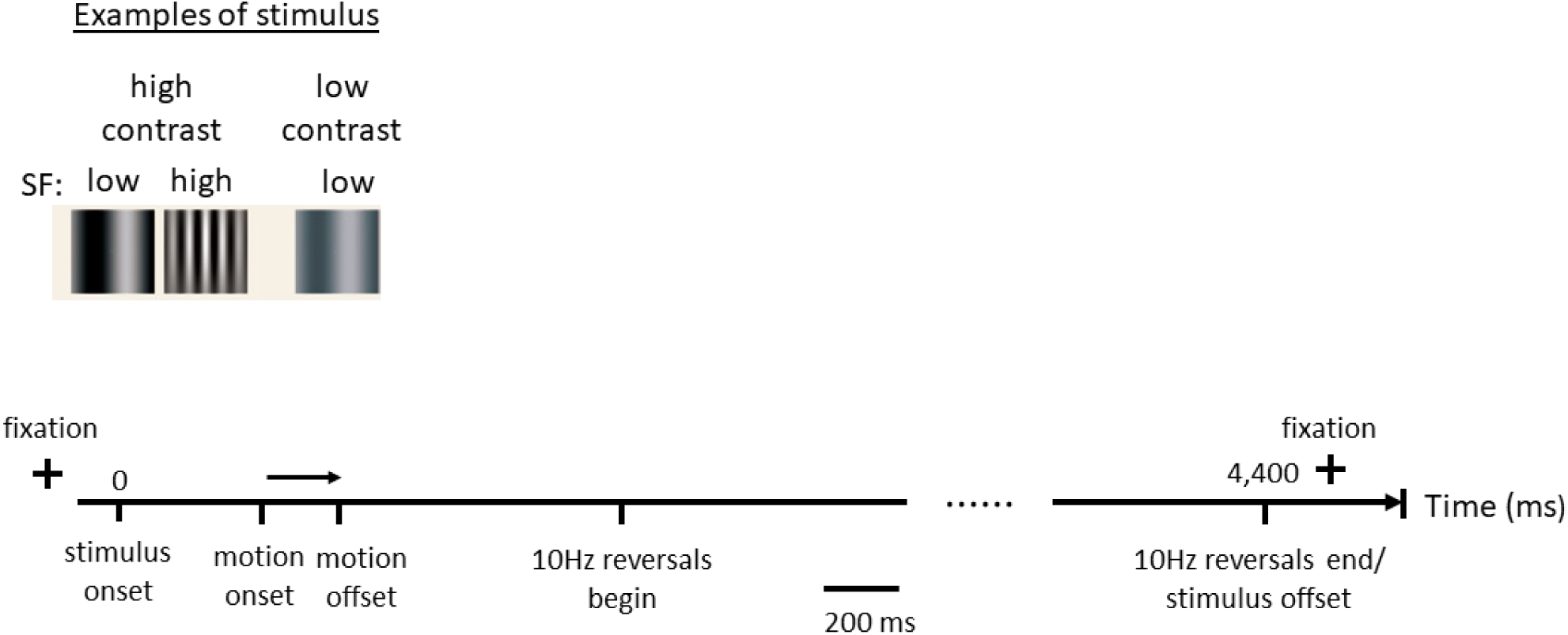
Schematic diagram of the visual stimulation paradigm. Stimuli varied in spatial frequency (SF) and contrast, with 10 Hz counter phase reversals to generate steady-state visual evoked potentials.

### Automated EEG preprocessing

Collected EEG data were processed using an automated preprocessing pipeline developed in Python, primarily utilizing the MNE library for EEG data analysis. The preprocessing pipeline involved several steps: (1) applying a notch filter (Comb IIR) at 50 Hz and a band-pass filter (Butterworth IIR, 4th order) between 1 and 40 Hz; (2) resampling the EEG data to 256 Hz; (3) marking segments longer than 5 seconds without task-relevant markers as break (bad) segments; (4) identifying bad channels using the RANSAC method implemented in the PyPREP library and interpolating these channels using values from neighboring good channels; (5) detecting and correcting ocular artifacts, including blinks and eye movements, via Independent Component Analysis (ICA), treating the ’0Z’ channel as an electrooculogram reference; (6) identifying and correcting transient, large-amplitude (> 200 µV) artifacts through Principal Component Analysis (PCA); (7) re-referencing EEG signals to the average of all channels; and (8) segmenting the continuous EEG data into epochs from -0.5 to 4.4 seconds relative to stimulus onset, applying baseline correction using the -0.5 to 0 second interval, and discarding epochs exceeding 200 µV peak-to-peak amplitude. After preprocessing, epochs corresponding to each stimulus condition were averaged across trials for each EEG channel. To derive regional EEG measures, epochs were further averaged across groups of spatially adjacent channels. Finally, time-domain EEG metrics— such as event-related potentials, entrainment ratios, spectral connectivity, and entropy—were extracted from various brain regions and analyzed separately according to the specific visual stimuli that elicited them.

### Feature extraction

Specific analyses were conducted on the preprocessed epoch data. For spectral connectivity, the average coherence was calculated within the frequency range of 2–40 Hz. Spectral entropy was evaluated using Shannon entropy to quantify the complexity of the frequency distribution. The entrainment ratio was determined by calculating the power at 10 Hz relative to adjacent frequencies from 1.4 to 3.4 sec after trial onset. ERP analyses included computing the area under the curve (AUC) for P50, N/P100, N200, N300, etc. Additionally, the power of the time-frequency representation (TFR) was computed for windows around the peaks at 100 ms, 200 ms, and 300 ms. Spectral connectivity was analyzed across eight selected channel pairs spanning frontal, central, parietal, and occipital regions. In contrast, spectral entropy, entrainment ratio, and ERPs were assessed across channels within 18 predefined sub-regions (detailed in Appendix Table A.1).

### Feature Selection and Regression Modelling

We developed a feature selection and multivariate linear regression pipeline (Fig. 2) in Python, utilizing the MNE, SciPy, and scikit-learn libraries, to estimate PET-amyloid SUVR from neurophysiological EEG signals. Initially, extracted EEG features were evaluated, and those demonstrating correlations with the target variable (i.e., PET-amyloid SUVR, CSF *p*-tau181, FCSRT free recall score, or MMSE score) exceeding an optimized threshold (0.23–0.42, selected for best predictive performance) were retained. To minimize redundancy and multicollinearity, inter-correlations among the selected features were analyzed, and features were grouped if their inter-correlations surpassed a second optimized threshold (0.6–0.9). From each group, only the feature exhibiting the strongest correlation with the target variable was retained. After that, the dimensionality of the resulting selected feature set was further reduced using PCA. Participants were pseudo-randomly divided into training (80%) and test (20%) subsets, stratified based on diagnosis and PET-amyloid SUVR (for estimation of PET-amyloid SUVR, FCSRT free recall, and MMSE scores) or CSF *p*-tau181 levels (for estimation of CSF *p*-tau181) to ensure similar distributions across subsets. Within the training subset, the optimal number of principal components (PCs; 8–16 components) explaining 60–70% of the total variance was determined. A multivariate linear regression model was then developed based on these PCs, hyperparameters (e.g., alpha) were optimized, and the model’s predictive accuracy was assessed on the test subset. To enhance robustness and reduce the risk of overfitting, k-fold cross-validation (k = 5) was employed throughout the analysis.

**Figure 2.**
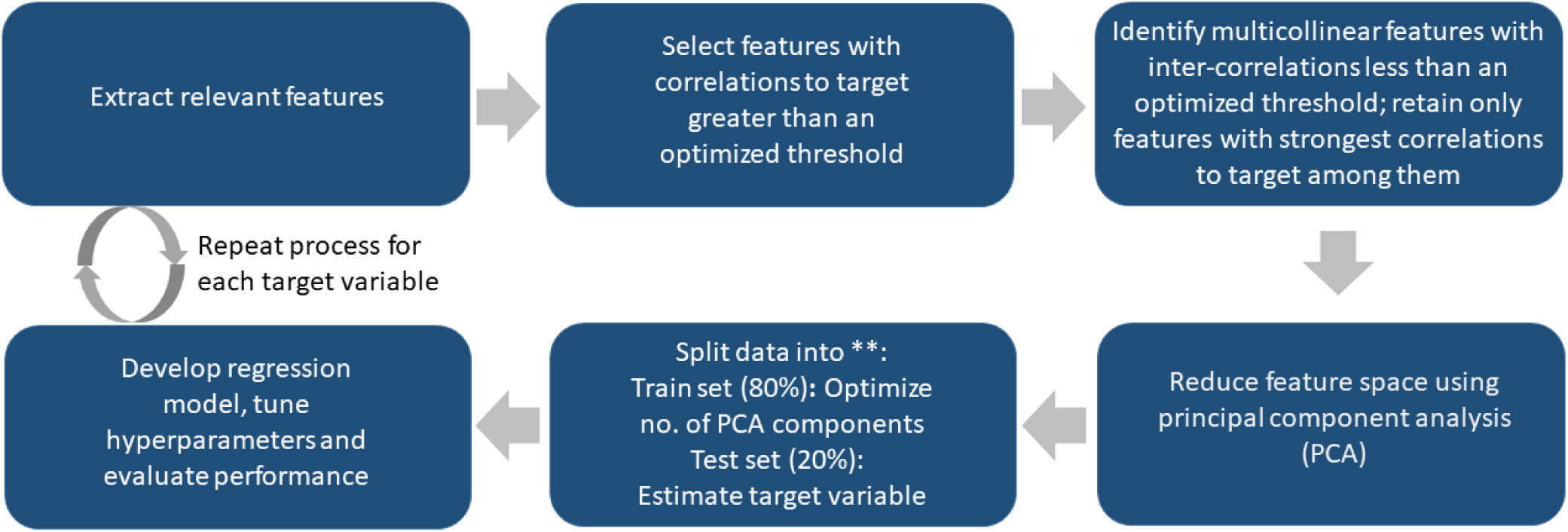
Feature selection and regression modeling pipeline. ** The train-test split was balanced and stratified according to participant diagnosis types and PET-amyloid SUVR. The model training and testing process was repeated across five different train-test splits to assess performance consistency.

## Results

### ERP amplitudes elicited by visual stimuli varied as a function of the diagnostic group

ERPs corresponding to the same stimulus condition were averaged across subsets of participants stratified by diagnostic group (control, MCI, and AD) or PET-amyloid SUVR level (high, mid, and low) to obtain grand average ERPs for each EEG channel. For instance, the grand average ERPs associated with one of the stimulus conditions were compared across different diagnostic groups (e.g., grand averages of 10 participants in each group shown in Fig. 3a) and PET-amyloid SUVR levels (e.g., grand averages of 16 participants in each level shown in Fig. 3b). Differences in ERP amplitude served as direct indicators of the functional integrity of neurons targeted by the respective stimulus. A trend was observed, showing reduced ERP amplitudes in participants with AD compared to controls, which was evident during the 0 to 0.4-sec time interval in Fig. 3a.

**Figure 3.**
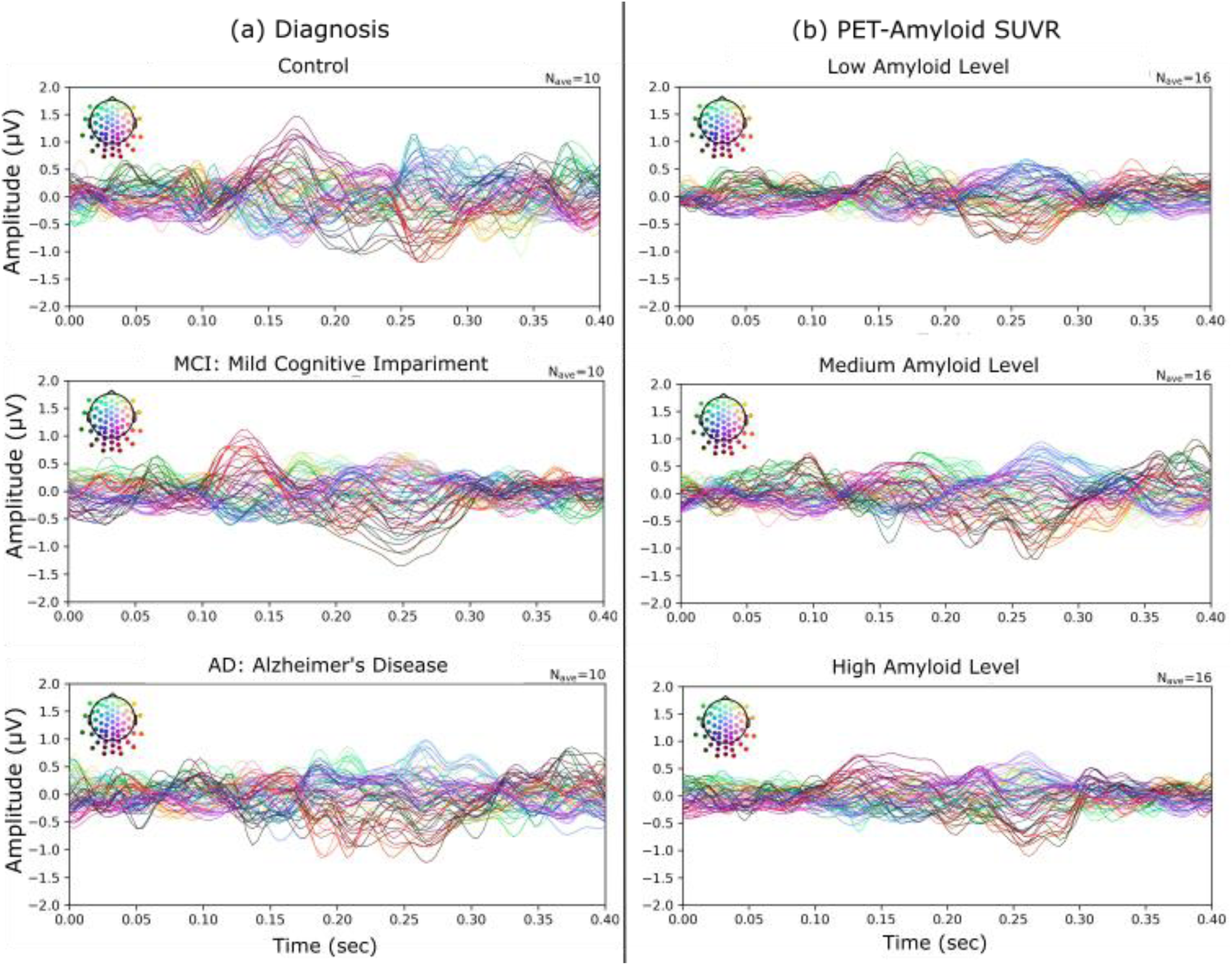
Grand average ERPs for the S2 stimulus, stratified by: (a) diagnosis (controls, MCI, AD; n=10 per group) and (b) PET-amyloid SUVR level (high, mid, low; n=16 per level).

### Regression modeling of selected ERP features estimated PET-amyloid SUVR with higher precision than CSF amyloid ratio

Five different train-test set splits were generated by stratifying based on the diagnostic group and PET-amyloid SUVR. Table 2 presents the training and test results obtained by repeating the regression modeling procedures on these 5 train-test splits. Correlations higher than 0.8 were observed in all train-test splits, with set 2 producing the best correlation coefficient. In set 2, a list of 243 features was identified using the feature selection method depicted in Fig. 2. PCA optimally reduced these selected features to 12 principal components (PCs), which collectively explained approximately 65% of the variance. The multivariate linear regression model was trained using these PCs and the true PET-amyloid SUVR from participants in the training set. This model was subsequently validated using the same 12 PCs in the test set to estimate PET-amyloid SUVR. Conversely, using shuffled PET-amyloid SUVR values in the training sets (repeated 50 times per train-test split), the resulting mean correlation coefficients for the combined training and test sets were consistently below 0.5 (Appendix Table A.2).

**Table 2.**
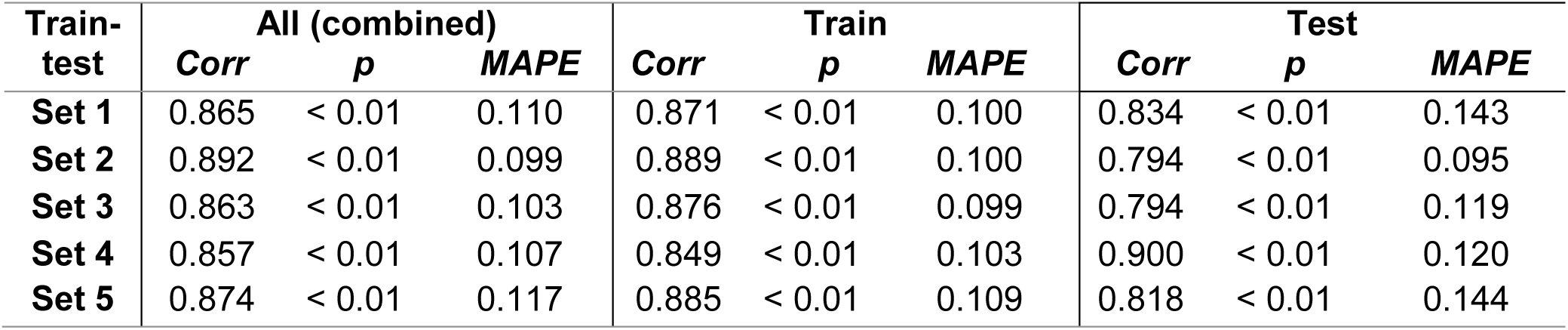
Correlation coefficient (*Corr*), *p*-values (*p*), and mean absolute percentage error (*MAPE*) across 5 different train-test splits for PET-amyloid SUVR.

Fig. 4a illustrates the Spearman correlation (r = 0.89, p < 0.01) between true and estimated PET-amyloid SUVR in the combined training and test sets for train-test set 2. For comparison, in Fig. 4b, the Spearman correlation between PET-amyloid SUVR and CSF amyloid ratio for all participants was -0.62 (p < 0.05). The absolute value of this correlation coefficient is lower than the correlation coefficient derived from the regression modeling. Hence, the PCs of the selected ERP features used in the regression model were better in estimating PET-amyloid SUVR than only using the CSF amyloid ratio as the predictor.

**Figure 4.**
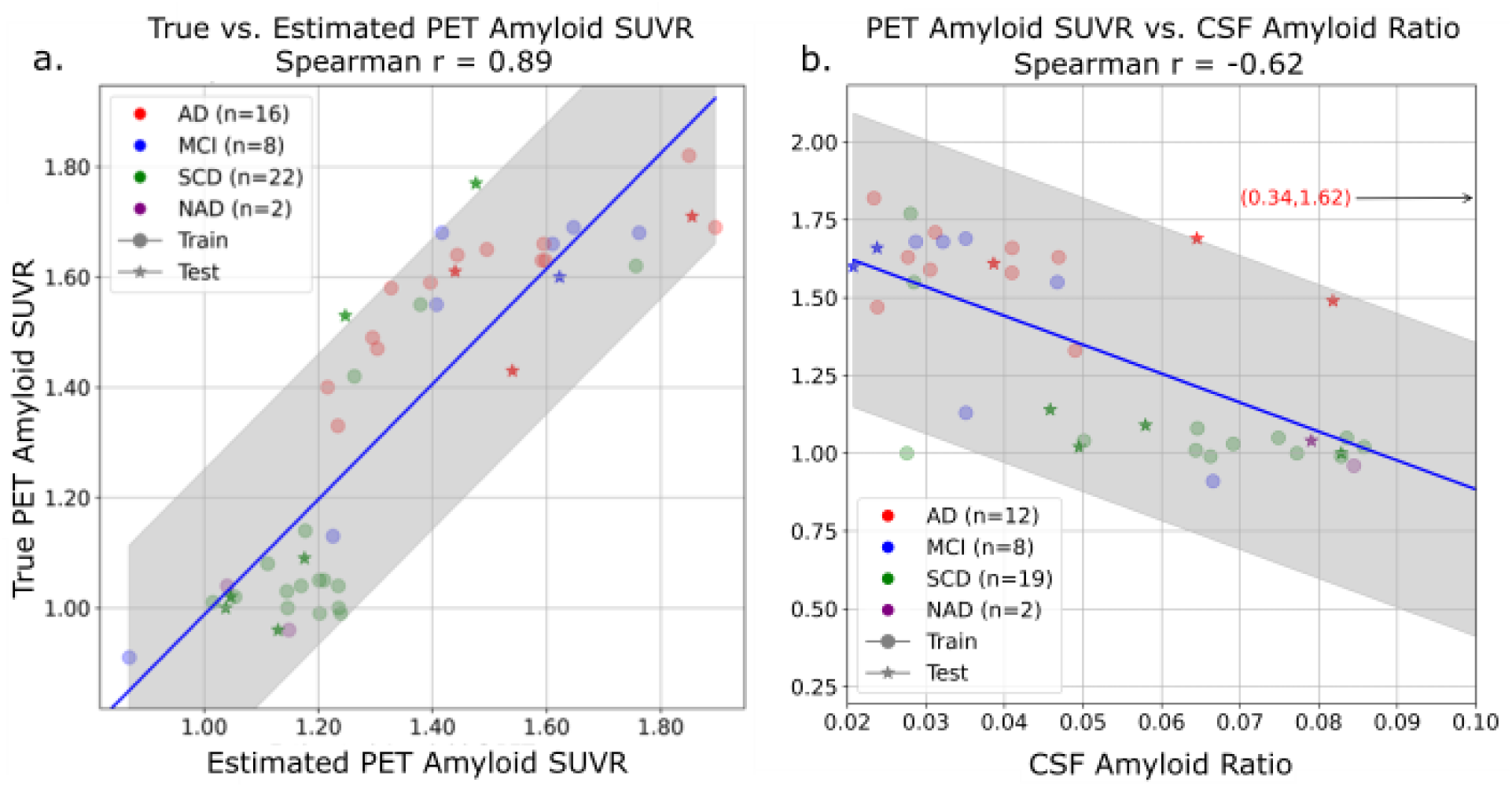
The regression model trained with the principal components generated from the selected ERP features performed better than the CSF amyloid ratio alone in estimating PET-amyloid SUVR. (a) Spearman correlation (r = 0.89, p < 0.01) between true and estimated PET-amyloid SUVR from the regression model developed using the train-test set 2 (Table 2). (b) Spearman correlation (r = -0.62, p < 0.05) between PET-amyloid SUVR and CSF amyloid ratio across all participants. Participants are labeled by diagnostic types using different colors and by train and test sets using different marker types. The coordinate labeled in red represents a significant outlier, omitted from the figure to preserve interpretability. The gray area indicates the 95% prediction interval.

### Strong correlations between true and estimated CSF p-tau181 levels using regression modeling of selected ERP features

We followed similar procedures, as illustrated in Fig. 2, to train and evaluate regression models to estimate CST *p*-tau181 levels. Participants were divided into training and testing datasets through five separate splits, stratified according to diagnostic groups and CSF *p*-tau181 levels. Table 3 summarizes the results from these regression analyses across all five train-test splits, with correlation coefficients ranging from 0.86 to 0.92. The best performance was achieved in set 3, yielding the highest correlation. In this train-test set, 202 relevant features were identified through the feature selection process. These features were subsequently reduced using PCA to 10 PCs, explaining approximately 64% of the total variance. A multivariate linear regression model was trained on these PCs using the participants’ true CSF *p*-tau181 values from the training set. The trained model was then validated on the test dataset using the same PCs to estimate CSF *p*-tau181 levels. Fig. 5 depicts the Spearman correlation (r = 0.92, p < 0.01) for the combined train and test sets, demonstrating strong predictive performance.

**Table 3.**
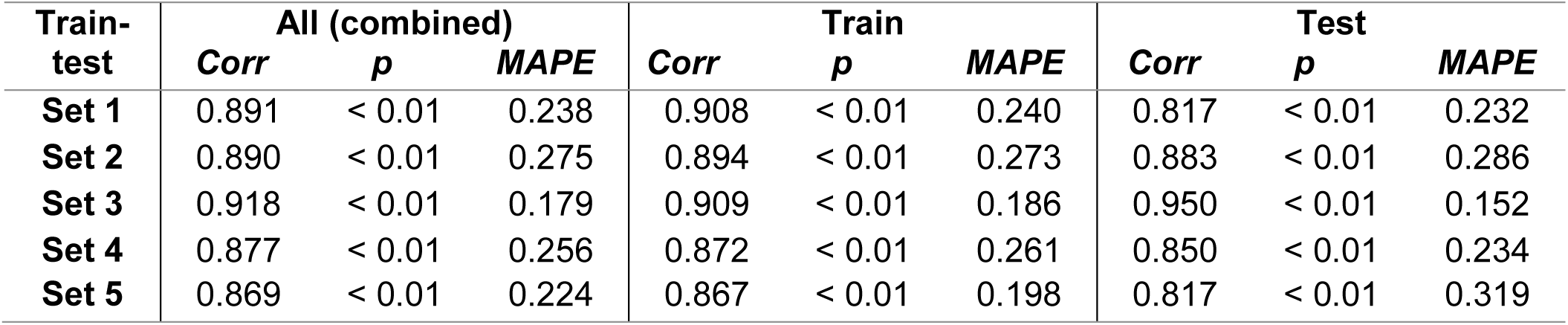
Correlation coefficient (*Corr*), *p*-values (*p*), and mean absolute percentage error (*MAPE*) across 5 different train-test splits for CSF *p*-tau181.

**Figure 5.**
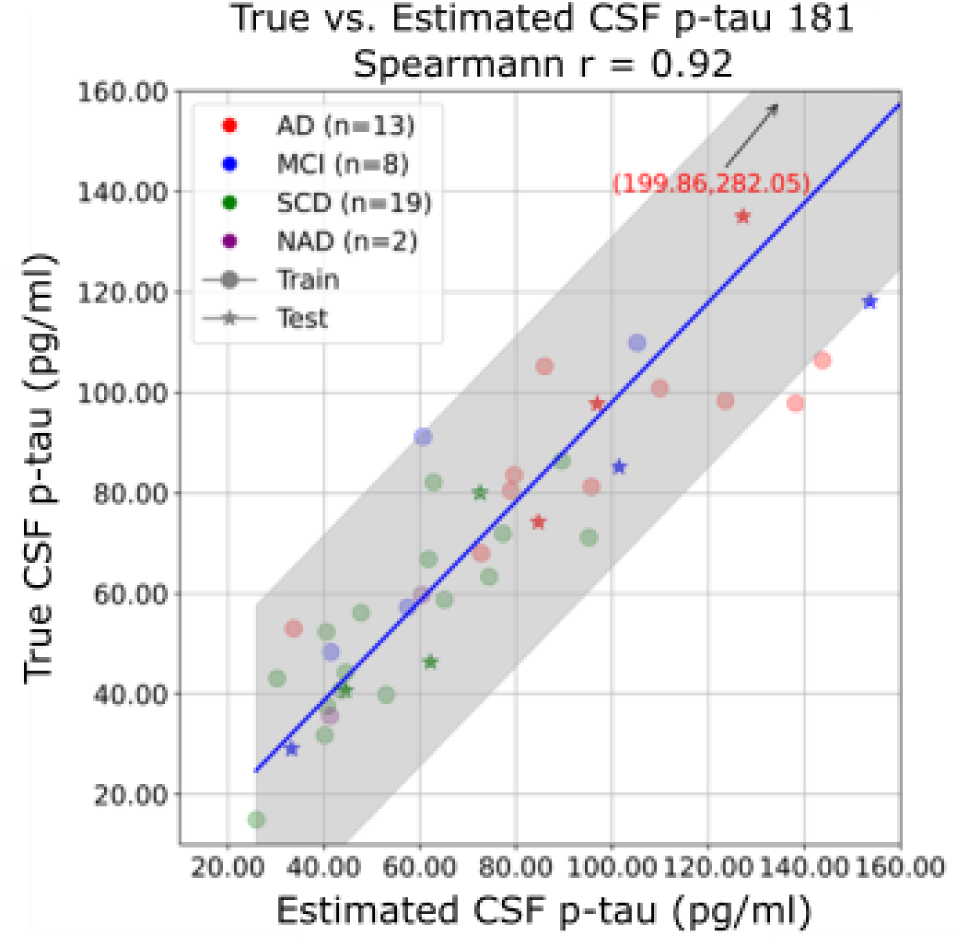
Spearman correlation (r = 0.92, p< 0.01) between true and estimated CSF *p*-tau181 levels from the regression model developed using the train-test set 3 (Table 3). Participants are labeled by diagnostic types using different colors and by train and test sets using different marker types. The coordinate labeled in red represents a significant outlier, omitted from the figure to preserve interpretability. The gray area indicates the 95% prediction interval.

### Estimation of Cognitive Performance (FCSRT Free Recall and MMSE Scores) via Regression Modeling of ERP Features

We applied similar procedures to train and evaluate multivariate linear regression models for estimating the FCSRT free recall and MMSE scores. Utilizing the same five train-test splits as used for PET-amyloid SUVR estimations, the results for the regression models are summarized in Tables 4 (FCSRT free recall) and 5 (MMSE). For the FCSRT free recall scores, the highest correlation coefficient was achieved in set 1. In this split, feature selection identified 338 relevant features, which were subsequently reduced into 11 PCs using PCA, collectively explaining approximately 66% of the variance. Similar to PET-amyloid SUVR estimations, a multivariate linear regression model was developed using these PCs along with the true FCSRT free recall scores from the training participants and was subsequently validated on the test participants using the same set of PCs to estimate their FCSRT free recall scores. Fig. 6a illustrates the Spearman correlation (r = 0.94, *p* < 0.01) for the combined training and testing datasets, demonstrating robust predictive performance.

**Figure 6.**
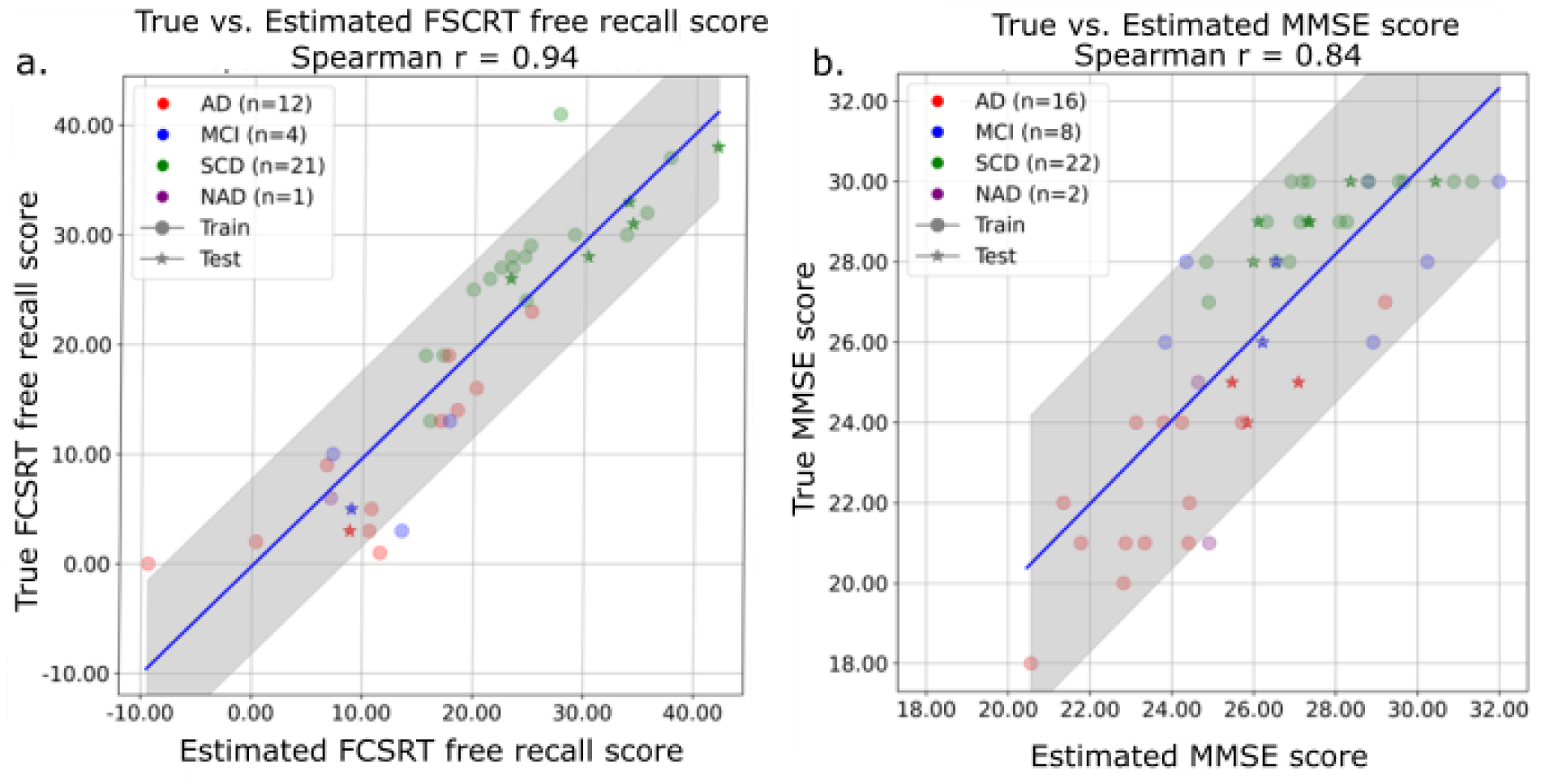
Estimation of FCSRT free recall and MMSE scores. (a) Spearman correlation (r = 0.94, *p* < 0.01) between true and estimated FCSRT free recall scores from the regression model developed using the train-test set 1 (Table 4). (b) Spearman correlation (r = 0.83, p< 0.01) between true and estimated MMSE scores from the regression model developed using the train-test set 1 (Table 5). Participants are labeled by diagnostic types using different colors and by train and test sets using different marker types. The gray area indicates the 95% prediction interval.

**Table 4.**
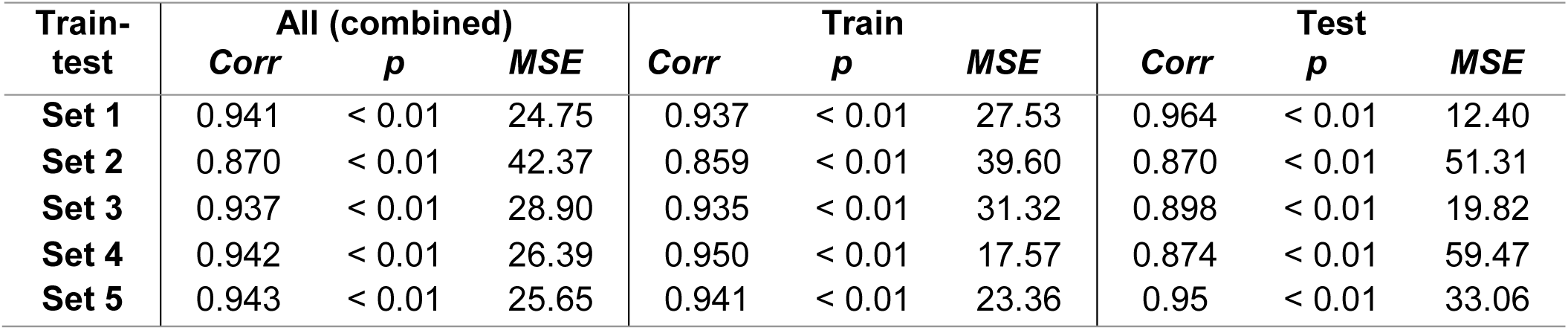
Correlation coefficient (*Corr*), *p*-values (*p*), and mean squared error (*MSE*) across 5 different train-test splits for FCSRT free recall.

**Table 5.**
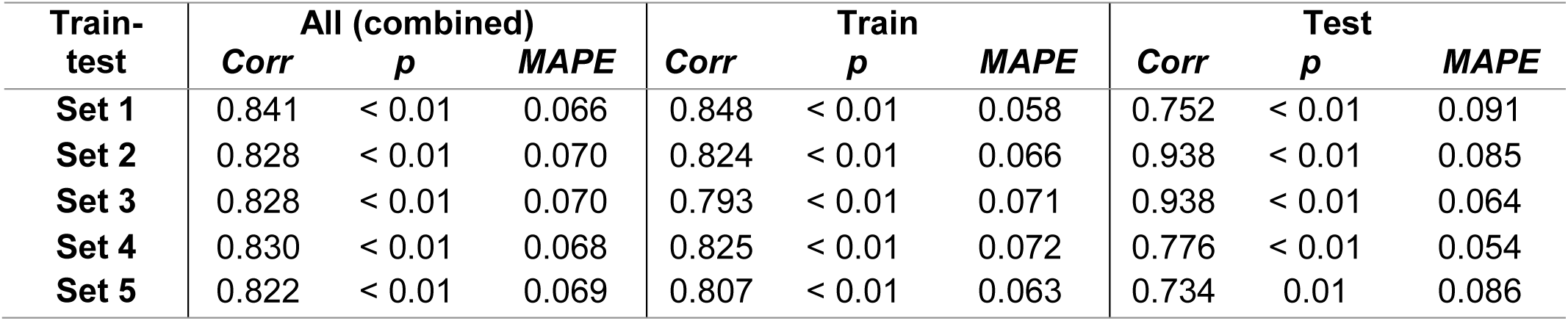
Correlation coefficient (*Corr*), p-values (*p*), and mean absolute percentage error (*MAPE*) across 5 different train-test splits for MMSE.

Meanwhile, for the MMSE scores, the highest correlation coefficient was again observed in set 1. In this set, the feature selection procedure yielded 170 relevant features, which were then reduced to 9 PCs via PCA, explaining approximately 60% of the total variance. A multivariate linear regression model trained with these PCs and true MMSE scores from the training participants was evaluated on the test set. Fig. 6b demonstrates the Spearman correlation (r = 0.84, p < 0.01) of the combined train and test sets.

## DISCUSSION

Healthcare providers face significant challenges in effectively treating dementia, largely because common initial symptoms often mask a spectrum of diverse, progressive diseases that are difficult to differentiate, monitor, and specifically address. To tackle these difficulties, the Amyloid, Tau, Neurodegeneration (ATN) diagnostic framework was established to enhance AD detection and monitoring [33]. Nevertheless, the practical implementation of the ATN framework and similar diagnostic models remains challenging due to the absence of a single portable technology capable of simultaneously assessing all necessary biomarkers.

One promising early indicator of AD pathology is altered neural activity. In humans, combined MRI and PET imaging studies have demonstrated correlations among cortical Aβ accumulation, early synaptic loss, and changes in global functional connectivity occurring before cognitive symptom onset [12]. Resting-state EEG analyses have also identified similar cortical network alterations linked to AD. However, despite these insights, resting-state EEG measures currently lack the sensitivity and specificity required for widespread adoption as screening or clinical decision-support tools.

To address these limitations, the present study utilizes the EPT protocol coupled with an automated pipeline. This protocol facilitates sensitive interneuron evaluation by employing targeted, sequential stimulations across spatially distinct neural pathways [23,34], thereby generating a functional map of interneuron integrity. Building upon previous research demonstrating EEG’s predictive capability for amyloid positivity using this protocol, we integrated it with an automated EEG preprocessing and feature extraction pipeline to estimate PET-amyloid SUVR and other related biomarkers of AD (i.e., CSF *p*-tau181, FCSRT free recall, and MMSE scores). Our study validated that this integrated approach effectively identifies pathophysiological brain changes in AD patients and control participants.

Specifically, the pipeline estimated PET-amyloid SUVR values with strong correlations (r > 0.8) between actual and predicted values, surpassing the predictive performance of CSF amyloid ratios alone. Previous research by Wisch et al. (2022) estimated continuous PET-amyloid values from CSF amyloid ratios using linear and generalized additive models, resulting in mean absolute percentage errors (MAPEs) of 23.8% and 15.1%, respectively [35]. In contrast, the best MAPE that we achieved using our regression modeling pipeline was 10%. Additionally, applying the same procedures and pipeline to estimate CSF *p*-tau181 levels, Free FCSRT free recall scores, and MMSE scores resulted in similarly robust correlations (0.8– 0.94) with their true values. These findings demonstrate that the integration of the EPT protocol and our automated pipeline provides a powerful method that enables the estimation of multiple important AD biomarkers.

However, this study’s findings are constrained by several limitations, including its modest sample size of 38-48 participants (depending on the target variable) and cross-sectional design, which restricts statistical power and longitudinal analyses. Additionally, due to the invasive nature of the clinical assessments required, no healthy controls were explicitly recruited. The participants represented a biased cohort with significant cognitive concerns warranting clinical PET scans, thus having a higher likelihood of elevated amyloid levels sufficient to explain cognitive deficits. This cohort bias restricts the generalizability of the findings and limits conclusions regarding potential benefits for population-wide screening. Moreover, the necessary exclusion criteria (e.g., older adults without complex neurological histories, such as prior strokes) further limit the general applicability of these findings. Despite these limitations, the results from this clinical study strongly suggest that key diagnostic criteria for AD can be reliably estimated using a single 30-minute EEG session.

AD remains a critical global health challenge, emphasizing the urgent need for reliable, portable, and non-invasive methods for early detection and disease staging. Identifying AD pathophysiology prior to the onset of cognitive symptoms could provide valuable opportunities for initiating preventative interventions. Altered neural activity, observable via techniques such as EEG and ERPs, holds substantial promise as an early non-invasive biomarker for AD. Specifically, EPT demonstrated an effective correlation between neural features and multiple AD biomarkers. Given EEG’s portability and non-invasive nature, this EPT protocol may effectively meet global demands for reliable AD screening and diagnostic tools. Further research and validation of this protocol as a screening methodology could significantly enhance clinical care and research in AD, enabling earlier detection, more precise cohort selection, and targeted therapeutic interventions.

## Abbreviations

AD: Alzheimer’s disease
ATN: Amyloid, Tau, Neurodegeneration
AUC: Area under the curve
Aβ: Amyloid-β
CERAD: Consortium to Establish a Registry for Alzheimer’s Disease Neuropsychological Battery
CSF: Cerebrospinal fluid
EEG: Electroencephalography
ELISAs: Enzyme-linked immunosorbent assays
EPT: Evoked Potential Tomography
ERP: Event-related potential
FCSRT: Free and Cue Selecting Reminding Test
LMU: Ludwig-Maximilians-University
MCI: Mild cognitive impairment
MMSE: Mini-Mental State Examination
MRI: Magnetic resonance imaging
NAD: Non-Alzheimer’s dementia
PCA: Principal component analysis
PET: Positron-emission-tomography
p-tau: Phosphorylated tau
SCD: Subjective cognitive decline
SUVR: Standardized uptake value ratio
TFR: Time-frequency representation

## Acknowledgments

R.P. was supported by the German Center for Neurodegenerative Disorders (Deutsches Zentrum for Neurodegenerative Erkrankungen, DZNE), the Hirnliga e.V. (Manfred-Strohscheer Stiftung), the Davos Alzheimer’s Collaborative, the VERUM Foundation, the Robert-Vogel-Foundation, the German Center for Neurodegenerative Diseases (DZNE), the National Institute for Health and Care Research (NIHR) Sheffield Biomedical Research Centre (NIHR203321), the University of Cambridge - Ludwig-Maximilians-University Munich Strategic Partnership within the framework of the German Excellence Initiative and Excellence Strategy and the European Commission under the Innovative Health Initiative program (project 101132356). R.P. and G.H. were supported by the Deutsche Forschungsgemeinschaft (DFG, 1007 German Research Foundation) under Germany’s Excellence Strategy within the framework of 1008 the Munich Cluster for Systems Neurology (EXC 2145 SyNergy - ID 390857198). B.S. and M.B. were supported by the Hirnliga e.V. (Manfred-Strohscheer Fellowship) for the ActiGlia project. G.H. was supported by the JPND Consortium SynOD “alpha-Synuclein OMICS to identify Drug-targets” (funded by the Federal Ministry of Education and Research, BMBF, 01ED2405A).

## Conflicts of interest

R.P. has received honoraria for advisory boards and speaker engagements from Roche, EISAI, Eli Lilly, Biogen, Janssen-Cilag, AstraZeneca, Schwabe, Grifols, Novo Nordisk, and Tabuk. Vistim Labs, Inc. provided personnel for this research on EEG-based diagnostics for dementia. The results have potential commercial implications for Vistim Labs. M.B. is a member of the Neuroimaging Committee of the EANM. M.B. has received speaker honoraria from Roche, GE Healthcare, Iba, and Life Molecular Imaging; has advised Life Molecular Imaging and GE Healthcare; and is currently on the advisory board of MIAC. N.F. has received honoraria for consulting and speaker engagements from GE Healthcare, Life Molecular Imaging, EISAI, Biogen, and MSD.

## Data Availability Statement

All data produced in the present study are available upon reasonable request to the authors.

## Appendix

**Table A. 1:** Eighteen defined sub-regions and their corresponding channel names in event response potential analysis.

| Sub-regions | Name of Channels |
| --- | --- |
| Frontal | 1Z, 2Z, 3Z, 2L, 2R, 3L, 3R |
| Central | 3Z, 4Z, 5Z, 4L, 4R, 5L, 5R |
| Parietal | 5Z, 6Z, 7Z, 6L, 6R, 7L, 7R |
| Occipital | 7Z, 8Z, 9Z, 8L, 8R, 9L, 9R, 10L, 10R |
| Right Frontal | 1RA, 1RB, 1RC, 2RB |
| Left Frontal | 1LA, 1LB, 1LC, 2LB |
| Right Central | 4R, 5R |
| Left Central | 4L, 5L |
| Right Parietal | 6R, 7R |
| Left Parietal | 6L, 7L |
| Right Occipital | 8R, 9R, 10R |
| Left Occipital | 8L, 9L, 10L |
| Right Anterior Temporal | 1RD, 2RC, 2RD |
| Left Anterior Temporal | 1LD, 2LC, 2LD |
| Right Medial Temporal | 2RA, 3RA, 3RB, 4RB, 3RC |
| Left Medial Temporal | 2LA, 3LA, 3LB, 4LB, 3LC |
| Right Parietal Temporal | 5RB, 4RC, 5RC, 3RD, 4RD |
| Left Parietal Temporal | 5LB, 4LC, 5LC, 3LD, 4LD |

**Table A. 2.** Correlation coefficients (Corr), *p*-values (*p*), and mean absolute percentage errors (MAPE) for 5 train-test splits using shuffled PET-amyloid SUVR values in the training sets.

| Train-test | All (combined) |  |  | Train |  |  | Test |  |  |
| --- | --- | --- | --- | --- | --- | --- | --- | --- | --- |
|  | *Corr* | *p* | *MAPE* | *Corr* | *p* | *MAPE* | *Corr* | *p* | *MAPE* |
| **Set 1** | 0.453 | < 0.01 | 0.176 | 0.553 | < 0.01 | 0.168 | -0.050 | 0.342 | 0.205 |
| **Set 2** | 0.426 | 0.016 | 0.178 | 0.504 | < 0.01 | 0.167 | 0.061 | 0.270 | 0.221 |
| **Set 3** | 0.450 | 0.019 | 0.176 | 0.556 | < 0.01 | 0.162 | -0.016 | 0.349 | 0.223 |
| **Set 4** | 0.434 | 0.013 | 0.175 | 0.531 | < 0.01 | 0.164 | -0.057 | 0.459 | 0.214 |
| **Set 5** | 0.450 | 0.018 | 0.179 | 0.540 | < 0.01 | 0.167 | 0.046 | 0.296 | 0.217 |

## Notes

### Author Declarations

Local Ethics committee of Ludwig-Maximilians-University (LMU) Munich gave ethical approval for this work (project numbers 17-755 and 17-569).

